# Forecasting left ventricular systolic dysfunction in heart failure with artificial intelligence

**DOI:** 10.1101/2025.04.13.25325744

**Authors:** Teya Bergamaschi, Tiffany Yau, Payal Chandak, Abena Kyereme-Tuah, Judy Hung, Hanna Gaggin, Isaac S. Kohane, Collin M. Stultz

## Abstract

**Background:** Objective assessment of left ventricular function remains a key prognosticator that is used to guide therapeutic decisions for patients with heart failure (HF). However, the left ventricular ejection fraction (LVEF) is dynamic, with worsening LVEF linked to increased morbidity and mortality. Identifying patients at risk of LVEF decline would improve prognostication and enable timely therapeutic intervention.

**Methods:** We developed a deep learning model to <u>P</u>redict changes in left ventric<u>UL</u>ar <u>S</u>ystolic function from <u>E</u>lectrocardiograms (ECG) of patients who have <u>H</u>eart <u>F</u>ailure (PULSE-HF). The model integrates 12-lead ECG waveforms with a patient’s history of prior LVEF measurements to calculate the likelihood that the LVEF will be less than 40% during the year after the ECG is obtained. The model is retrospectively developed and tested using data from one hospital and externally validated on retrospective cohorts from two different hospitals.

**Findings:** PULSE–HF demonstrates strong discriminatory ability with respect to forecasting whether the LVEF would be below 40% within the next year, achieving areas under the receiver operating characteristic curve (AUROC) of 87.5-91.4% across all three HF cohorts. Among patients with HF who have a baseline LVEF above 40%, PULSE–HF effectively identified those at risk of worsening LVEF with AUROCs of 81.6-86.3% across all three datasets. PULSE–HF’s discriminatory ability remained consistently high across a range of subgroups with different comorbidities and regardless of medical therapy. Assuming an underlying prevalence of LVEF worsening of 10% per year, PULSE-HF’s negative predictive values are over 97%, assuming an underlying sensitivity of 80%. Lastly, we demonstrate that a lead I version of PULSE–HF has a performance similar to the performance of the model that uses all 12 ECG leads.

**Interpretation:** PULSE–HF robustly predicts worsening LVEF in patients who have a prior diagnosis of HF. The method provides a platform for identifying patients who are at an increased risk of worsening systolic dysfunction.

**Funding:** This work was supported, in part, by a grant from Quanta Computers.

**Research in context:** *Evidence before this study:* Heart failure (HF) remains a leading cause of morbidity and mortality across the globe. Left ventricular ejection fraction (LVEF) plays an important role in prognostication and treatment of HF. Moreover, LVEF is dynamic, with worsening LVEF being associated with increased mortality and poor outcomes compared to patients with HF with stable LVEF. Identifying patients with HF at risk of future decline in LVEF would improve prognostication and facilitate the timely delivery of appropriate medical therapy. However, models that use the electronic health record (EHR) to forecast declines in LVEF have demonstrated modest discriminatory ability. For instance, a model incorporating echocardiographic features, demographics, and comorbidities from the EHR had areas under the receiver operating characteristic curve (AUROC) between 54%–68% for predicting a 30% decline in LVEF within one year after HF diagnosis. Electrocardiograms (ECG) may contain information about LVEF, as deep learning models of ECGs have been used to estimate the contemporaneous LVEF. A prior method uses ECG to forecast LVEF below 40%, but this method was developed for a much shorter future time window of six months and specifically for patients with premature ventricular complexes. To date, the ECG has not been used to forecast future changes in LVEF among HF patients. Overall, no robust method exists for identifying patients with HF at high risk of future LVEF decline.

*Added value of this study:* We developed a deep learning model, called PULSE–HF, to <u>P</u>redict changes in left ventric<u>UL</u>ar <u>S</u>ystolic function from <u>E</u>CGs of patients who have <u>H</u>eart <u>F</u>ailure. The model leverages both the 12-lead ECG and history of prior LVEF measurements to identify patients at risk of having a reduced LVEF (≤ 40%) within the year after the ECG was obtained. We also developed a single lead version of PULSE-HF, based on lead I, that maintains strong performance compared to the model that uses all 12 leads.

*Implications of all the available evidence:* The ECG contains information that can be leveraged to forecast changes in LVEF. As ECGs are typically obtained for patients with HF during yearly follow-up visits, we envision that the method could be used annually to identify patients who are at the highest risk of future LVEF decline. Screening could be routinely integrated into existing clinical workflow with minimal changes to clinical practice. Since similar performance was achieved with a single lead model (lead l), it may be possible to complete screening using wearable or pocket ECG monitoring devices. Prospective studies are needed to better define the utility of PULSE–HF in guiding HF management decisions, optimizing prediction windows, and integrating the model into clinical practice.

## Introduction

Heart failure (HF) remains a leading cause of morbidity and mortality in individuals over 65 years of age (1). Despite advances in diagnosis and treatment, five-year mortality rates in patients with HF exceed 50% in many cohorts, indicating the need for improved risk stratification and therapies (1). A cornerstone of HF management is an objective assessment of left ventricular systolic function, typically quantified as the left ventricular ejection fraction (LVEF). The LVEF is used to classify HF into three mutually exclusive subtypes: HF with reduced ejection fraction (HFrEF, LVEF ≤ 40%); HF with mildly reduced ejection fraction (HFmrEF, LVEF 41–49%); and HF with preserved ejection fraction (HFpEF, LVEF ≥ 50%) (2). Therapeutic decision making is a function of the subtype that a patient belongs to, e.g., guideline directed medical therapy (GDMT) for patients with HFrEF has been shown to reduce mortality by 73% over two years, and current recommendations for initiating quadruple therapy are largely a function of this dichotomous cutoff (LVEF< 40%) (3). Moreover, LVEF plays an important role in prognostication, as decreases in LVEF below 45% are associated with proportional increases in the hazard ratio for all-cause mortality (4).

LVEF is dynamic and may increase with appropriate therapy or decrease as HF progresses over time (5, 6). For example, longitudinal studies show that about one out of three patients with HFpEF experience a decline in their LVEF to below 40% within two years of diagnosis (7,9). Indeed, worsening LVEF has been associated with an approximately two-fold increase in mortality relative to stable HFpEF diagnosis, as well as worse outcomes compared to a stable HFrEF diagnosis (8–11). A method that accurately identifies patients with HF at high risk of LVEF decline would improve prognostication, optimize echocardiogram scheduling, and enable timely initiation of GDMT.

Although a number of studies have found clinical features that are correlated with decreasing LVEF, risk models that utilize these clinical features to forecast a significant decline in LVEF have shown modest discriminatory ability (9,11). For instance, an extreme gradient boosting (XGBoost) model incorporating electronic health record (EHR) data––including echocardiographic features, demographics, and comorbidities––had areas under the receiver operating characteristic curve (AUROC) between 54%–68% for predicting a 30% decline in LVEF within one year after HF diagnosis (12). Collectively, the modest results from tabular EHR-based models suggest that tabular EHR data, including features known to be correlated with LVEF decline, have limited predictive value for the task of forecasting declines in LVEF.

Prior studies suggest that deep learning models using the electrocardiogram (ECG) can identify subclinical markers of systolic dysfunction not captured in structured EHR data (13,14). However, most of these models use the ECG to classify the concurrent LVEF rather than forecasting LVEF decline (13,14). As such, these approaches rely on LVEF measurements associated with echocardiograms that are acquired at a time close to when the ECG is obtained and consequently are not designed to identify patients who will have a significant decline in their LVEF in the future. Moreover, these models were not specifically developed on patients who have HF. One prior method forecasted reduced LVEF, but it was developed specifically for patients who have premature ventricular complexes, rather than those with HF, and for a much shorter future period of 6 months (15).

We propose a deep learning model, called PULSE–HF, to <u>P</u>redict changes in left ventric<u>UL</u>ar <u>S</u>ystolic function from <u>E</u>CGs of patients who have <u>H</u>eart <u>F</u>ailure. PULSE–HF integrates 12-lead ECG waveforms with LVEF information from echocardiograms obtained before the ECG is acquired (i.e., the LVEF history), to forecast the likelihood that a patient’s LVEF will be less than 40% within one year after the ECG. We validate the model in retrospective HF cohorts from three different hospitals. Notably, among patients with HF with baseline LVEF above 40%, PULSE–HF effectively identifies those at risk of worsening LVEF in all three datasets. We also present a single-lead version of PULSE–HF (based on lead I) and find that its performance is similar to the 12-lead version. PULSE-HF provides a platform to identify patients with HF who are at increased risk of worsening systolic dysfunction.

## Methods

### Study design

PULSE-HF was developed and validated using clinical data from three institutions: Massachusetts General Hospital (Hospital 1), Brigham and Women’s Hospital (Hospital 2), and a deidentified-dataset from PhysioNet (MIMIC IV, https://physionet.org/content/mimic-iv-ecg/1.0/). In all cohorts, only patients with a confirmed HF diagnosis, based on ICD codes, and who had at least one ECG and one LVEF measurement were included. These cohorts were further refined to include only those patients who had at least one LVEF within a year following an ECG, so a label could be defined. The study population from Hospitals 1 and 2 included ECGs collected between January 1, 2000, and June 30, 2021, and the study population from MIMIC IV included ECGs collected between 2008 and 2019. Data from Hospital 1 were used to develop and test the model, and data from the other two institutions were used to externally validate the approach. This retrospective study was approved by MGB Institutional Review Board protocol number 2020P000132, with a waiver of informed consent as the data were obtained from routine clinical care.

### Cohort characteristics

PULSE–HF was evaluated across three institutions; cohort characteristics across demographics, comorbidities, and medications are summarized in Table 1. At Hospital 1, the model was validated on an internal holdout dataset of 65,908 ECGs from 7,008 patients. At Hospital 2 and MIMIC, the model was validated on external holdout datasets of 106,475 ECGs from 13,211 patients and 65,558 ECGs from 8,835 patients, respectively. Patients had an average of 3.8, 2.3, and 2.3 years, and up to 20.7, 21.3, and 10.9 years of ECG records post HF diagnosis in Hospital 1, Hospital 2, and MIMIC, respectively. The cohorts had statistically significant differences in age, sex, LVEF distribution, comorbidity diagnosis, and medication prescription. For patients at Hospital 1, echocardiograms were examined per patient at the time of diagnosis and the following prevalence estimates were observed: left atrial enlargement (≥ mild: 6.4%, ≥ moderate: 4.0%), abnormal LV wall thickness (5.8%), LV hypertrophy (25.9%), any mitral regurgitation (3.3%; ≥ moderate: 2.0%), and aortic stenosis (4.3%).

**Table 1.** Cohort characteristics.

|  | Hospital 1 | Hospital 2 | MIMIC |
| --- | --- | --- | --- |
| Patients, n | 46,694 (39,686 development;<br>7,008 test) | 13,211 | 8,835 |
| Sex, % (n) | 42.6 (19,879) | 44.7 (5,900) | 46.7 (4,129) |
| Age, mean ± std | 70.6 ± 15.2 | 69.9 ± 14.4 | 73.1 ± 13.7 |
| ECGs, n | 426,081 | 106,475 | 65,558 |
| Prior 1-year LVEF support, mean | 1.10 | 1.07 | 0.96 |
| LVEF measurements, n | 179,979 | 35,549 | 23,209 |
| LVEF ≤ 40 (HFrEF), % (n) | 21.9 (39,348) | 35.3 (12,559) | 37.2 (8,624) |
| LVEF 41-49 (HFmrEF), % (n) | 9.4 (16,953) | 7.2 (2,559) | 6.7 (1,558) |
| LVEF ≥ 50 (HFpEF), % (n) | 68.7 (123,678) | 57.5 (20,431) | 56.1 (13,027) |
| Diabetes mellitus, % (n) | 42.7 (19,939) | 39.4 (5,200) | 39.4 (3,481) |
| Hypertension, % (n) | 83.6 (39,041) | 64.4 (8,508) | 61.0 (5,391) |
| Atherosclerotic heart disease % (n) | 67.2 (31,379) | 60.4 (7,982) | 56.8 (5,016) |
| Chronic obstructive pulmonary disease, % (n) | 13.6 (6,350) | 6.3 (828) | 4.8 (424) |
| Atrial fibrillation, % (n) | 53.3 (24,876) | 47.8 (6,320) | 51.4 (4,537) |
| Coronary artery bypass graft, % (n) | 7.7 (3,588) | 3.4 (443) | 6.9 (611) |
| Percutaneous coronary intervention, % (n) | 26.9 (12,578) | 22.4 (2,965) | 13.2 (1,162) |
| Myocardial infarction, % (n) | 43.7 (20,404) | 57.5 (7,590) | 27.9 (2,469) |
| ACEi/ARB, % (n) | 64.7 (30,231) | 50.8 (6,717) | 58.3 (5,154) |
| ARNi, % (n) | 9.3 (4,351) | 6.0 (795) | 8.7 (773) |
| Beta blockers, % (n) | 79.3 (37,005) | 64.8 (8,563) | 85.8 (7,579) |
| MRA, % (n) | 18.3 (8,543) | 10.6 (1,401) | 19.1 (1,688) |
| Diuretic, % (n) | 76.6 (35,787) | 77.0 (10,178) | 85.9 (7,590) |

Additionally, the ability of PULSE-HF to identify worsening LVEF in patients with a baseline above 40% was evaluated. Cohort characteristics across demographics, comorbidities, and medications among patients who had at least one LVEF above 40% after their HF diagnosis are summarized in Table 4. Clinical characteristics within this cohort were similar to the overall population at each corresponding site. Moreover, across the three clinical cohorts, approximately 17.4% of patients who begin with an LVEF over 40% eventually develop an LVEF below 40% over median and mean follow-up times of 25 and 42 months, respectively. The average rate of LVEF decline per year was about 8.1%, a finding similar to prior longitudinal studies, which found a prevalence of about 10% (11,12).

### Model development

PULSE-HF was designed to predict whether any LVEF measurements will be under 40% in the year after an ECG is obtained. Consequently, the task was formalized as a supervised learning problem where an ECG was labeled as positive if there was any echocardiogram that indicated an LVEF below 40% within the year after the date that the ECG was obtained, and negative if there was at least one LVEF measurement within the next year, but no LVEF measurements below 40% in that timeframe. Thus, if the LVEF was already below 40% when the initial ECG was acquired, then a positive PULSE-HF prediction corresponded to forecasting that the LVEF would remain below 40% in the following year. By contrast, if the preceding LVEF was above 40%, then a positive prediction for PULSE-HF forecasted that the LVEF would decline to below 40% in the subsequent year; i.e., that the LVEF would worsen.

Labeled ECGs from Hospital 1 were split by patient into a development set comprising 85% of the data and an internal test set comprising 15%. The development set was further divided into a training dataset that was used to learn model parameters, and a validation dataset that was used to select the best hyperparameters and model. ECGs from Hospital 2 and MIMIC served as external test datasets. All patients, including those who experienced rapid clinical deterioration (death, fatal arrhythmias, or HF hospitalization) or underwent advanced therapies such as heart transplantation, LVAD implantation, or CRT, were included in the dataset if all necessary data, i.e., an ECG and subsequent LVEF measurement, were available for such instances. All available LVEF measurements were included in the dataset; therefore, multiple ECG–LVEF pairs per patient were considered. Training, validation, and test were stratified by patient – ECG and LVEF data for one patient only appeared in one subset (either training, validation, or testing).

ECG waveforms across all cohorts were recorded for 10 seconds and sampled at a minimum of 250 Hz with amplitudes converted to millivolts. Linear interpolation was used to fit each lead into 2,500 samples and organize them into a 2,500 × 12 matrix with lead order {I, II, III, AVR, AVL, AVF, V1, V2, V3, V4, V5, V6}. ECGs were excluded if any sample exceeded 5 mV in absolute value, if any lead was entirely zero, or whose absolute value was below machine precision. If multiple ECGs were recorded within an hour, only the last one was retained.

In addition to the ECG waveform, features were generated using previously recorded LVEF measurements obtained from echocardiograms. If a patient had multiple echocardiograms recorded in their history, multiple LVEF measurements could be incorporated. More precisely, a patient’s the LVEF history includes the following metrics acquired before the ECG: the initial LVEF value at diagnosis of HF; the LVEF value immediately preceding the ECG; the count, mean, minimum, maximum, standard deviation, and range of LVEFs in the past year; whether any LVEFs under 40%, between 40–50%, and above 50% were measured in the past year; whether all LVEFs in the past year were classified as under 40%, between 40–50%, and above 50%; and the number of days since diagnosis of HF. If any of the LVEF history features were not available, these values were imputed with zeros.

PULSE-HF consists of a binary classification model using an ECG encoder (Figure S1A,B), an LVEF history encoder (Figure S2), and a prediction module (Figure S3). The ECG encoder consists of a ResNet-18 variant, with 12 input channels and 1D convolutions, used to produce an ECG embedding. A multilayer perceptron (MLP) encoder processes the LVEF history into another embedding. The ECG and LVEF history embeddings are summed, and the resulting vector is fed into an MLP module to generate a prediction. A schematic of the final model is shown in Figure S3. Single-lead versions of the PULSE–HF model were also trained, where the ECG encoder leverages only a single lead, and the rest of the model remains the same as the original PULSE-HF architecture.

Model parameters were optimized using a binary cross-entropy loss with 5% weight decay, an AdamW (16) optimizer, a batch size of 2048, and a learning rate scheduler with an initial learning rate of 10^-5^ and an end learning rate of 10^-8^. Dropout regularization with a probability of 0.3 and early stopping on the validation AUROC with patience of 20 epochs was applied to mitigate overfitting. Model development utilized PyTorch and PyTorch Lightning, with source code publicly available.

For comparison to a standard, non-deep learning-based method, a logistic regression model was trained using clinical features extracted from ECGs (Table S10) and the same summary statistics of LVEF history that were used in PULSE–HF. This model was developed for the same task as PULSE-HF; that is, to predict if any LVEF measurements will be under 40% in the year after an ECG is obtained. Logistic regression hyperparameters were optimized using Scikit-Learn by performing a grid search over the regularization strength with the range 1E-3 to 1E3 and testing both L1 and L2 regularization using the Stochastic Average Gradient Accelerated (SAGA) solver. The best hyperparameter combination was selected based on validation AUROC.

### Statistical analysis

Model performance was evaluated using AUROC. Sensitivity, specificity, positive predictive value (PPV), and negative predictive value (NPV) were computed across various operating points. Other longitudinal studies suggest that the prevalence of significant decrements in LVEF over one year is approximately 10% (11,12); therefore, predictive values were calculated for this underlying prevalence.

PULSE–HF was assessed on the internal test dataset at Hospital 1 and external test datasets at Hospital 2 and MIMIC using the best-performing model trained on the development set. Results are reported for various sensitivity thresholds determined on the training dataset. To evaluate robustness, performance was stratified across key clinical subgroups. Model performance was examined among ECGs with a baseline LVEF above 40% to determine its ability to forecast worsening LVEF. Single-lead versions of PULSE–HF were similarly evaluated to determine feasibility for remote monitoring. All 95% confidence intervals were estimated via 1000 bootstraps of the respective test datasets.

Saliency analysis was used to interpret the contributions of the different features in the Lead I PULSE-HF model (17,18). For each ECG, the absolute gradient of the loss function was computed for each input sample; i.e., a sample from the ECG or a LVEF history feature. For ECG saliency values, the temporal distribution of high-saliency points was analyzed by aligning these high-saliency points with R peaks detected via NeuroKit2 (19). Specifically, the most salient point in each ECG was identified, and its time distance to the next R peak was measured. For LVEF features, the mean absolute saliency value for each feature was measured and compared to the mean top-1 absolute saliency value from ECGs. All analyses were performed using Python.

## Results

PULSE-HF is designed to determine whether the LVEF will be below 40% within one year after an ECG is acquired using the ECG and previous LVEF measurements (Figure 1). The model is evaluated using retrospective data obtained from three hospitals.

**Figure 1.**
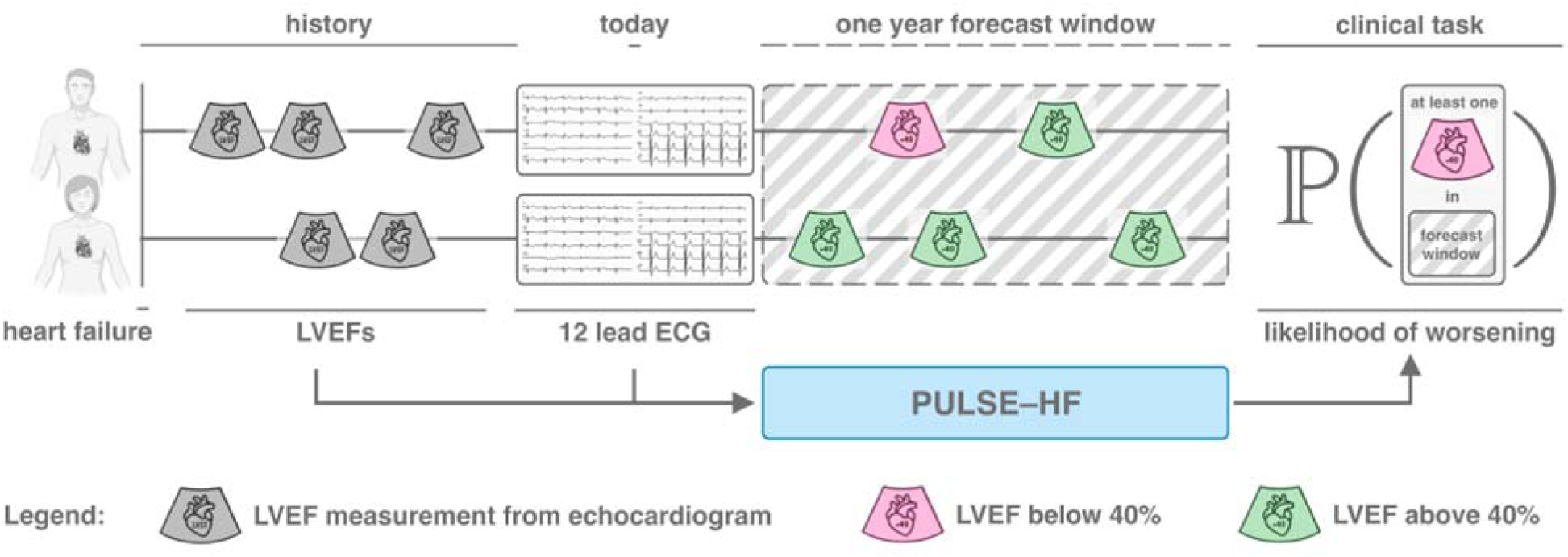
Overview of PULSE–HF. A deep learning model designed to Predict changes in left ventricULar Systolic function from Electrocardiograms of patients with HF. PULSE–HF integrates 12-lead ECGs with LVEF history to forecast the likelihood of LVEF falling below 40% within one year after the ECG.

### Forecasting low LVEF in adults with HF

The feasibility of accurately forecasting low future LVEF using ECG features and LVEF without deep learning was first explored through logistic regression modeling. A logistic regression model using summary ECG features (Table S10) developed on the same task achieved modest AUROCs (Hospital 1: 70.3%; Hospital 2: 69.2%). Incorporating the same LVEF history features used in PULSE–HF improved AUROCs (Hospital 1: 78.1%; Hospital 2: 74.1%).

PULSE–HF demonstrated strong discriminatory ability with respect to forecasting whether the LVEF would fall below 40% within the next year, achieving AUROCs of 87.5-91.4% across all three institutions (Table 2). When using ECGs alone, assuming no access to LVEF history, PULSE-HF maintained AUROCs of 79.1-84.0% across the three institutions. Overall model calibration was similarly good across all three institutions (Figure S9). Moreover, in all three cohorts, the discriminatory ability remained consistently high across a range of subgroups with different comorbidities and regardless of medical therapy (Table 2). Further, discriminatory ability remained strong in individual ECGs that exhibited atrial fibrillation or pacing in the Hospital 1 and Hospital 2 datasets (Table 3).

**Table 2.** Discriminatory ability for forecasting low LVEF in all patients, stratified by important clinical variables.

| Subgroup |  | AUROC |  |  |
| --- | --- | --- | --- | --- |
|  |  | Hospital 1 | Hospital 2 | MIMIC |
| All patients |  | 91.4 ± 0.2 | 87.6 ± 0.2 | 87.5 ± 0.3 |
| Average annual hospitalizations | < 1 | 91.0 ± 0.3 | 86.7 ± 0.3 | 84.5 ± 0.5 |
|  | ≥ 1 | 92.1 ± 0.4 | 89.4 ± 0.4 | 89.6 ± 0.3 |
| Diabetes mellitus | No | 92.0 ± 0.3 | 87.6 ± 0.3 | 86.7 ± 0.4 |
|  | Yes | 90.6 ± 0.4 | 87.5 ± 0.3 | 88.4 ± 0.4 |
| Hypertension | No | 90.9 ± 0.6 | 86.9 ± 0.4 | 86.6 ± 0.5 |
|  | Yes | 91.3 ± 0.3 | 87.5 ± 0.3 | 88.0 ± 0.3 |
| Atherosclerotic heart disease | No | 90.6 ± 0.5 | 87.0 ± 0.4 | 85.9 ± 0.5 |
|  | Yes | 91.5 ± 0.3 | 87.4 ± 0.3 | 87.7 ± 0.3 |
| Chronic obstructive pulmonary disease | No | 91.1 ± 0.3 | 87.4 ± 0.2 | 87.4 ± 0.3 |
|  | Yes | 92.9 ± 0.7 | 89.7 ± 0.7 | 89.4 ± 1.3 |
| Coronary artery bypass graft | No | 91.5 ± 0.3 | 87.4 ± 0.2 | 87.4 ± 0.3 |
|  | Yes | 89.1 ± 0.8 | 90.8 ± 0.9 | 87.0 ± 1.0 |
| Atrial fibrillation | No | 91.3 ± 0.4 | 88.2 ± 0.3 | 87.7 ± 0.4 |
|  | Yes | 91.5 ± 0.3 | 86.9 ± 0.3 | 87.3 ± 0.4 |
| ACEi/ARB | No | 89.4 ± 1.3 | 82.9 ± 0.9 | 89.2 ± 1.3 |
|  | Yes | 91.5 ± 0.2 | 87.9 ± 0.2 | 87.4 ± 0.3 |
| ARNi | No | 91.1 ± 0.5 | 86.3 ± 0.4 | 87.4 ± 0.5 |
|  | Yes | 91.4 ± 0.3 | 88.2 ± 0.3 | 87.0 ± 0.4 |
| MRA | No | 91.3 ± 0.3 | 87.3 ± 0.2 | 87.4 ± 0.3 |
|  | Yes | 92.2 ± 0.7 | 90.5 ± 0.7 | 88.4 ± 0.9 |
| Beta blockers | No | 89.8 ± 0.3 | 86.6 ± 0.2 | 85.6 ± 0.3 |
|  | Yes | 92.8 ± 0.4 | 90.9 ± 0.5 | 91.1 ± 0.5 |
| Diuretics | No | 90.1 ± 0.7 | 87.0 ± 0.4 | 89.3 ± 0.7 |
|  | Yes | 91.6 ± 0.2 | 87.8 ± 0.2 | 87.1 ± 0.3 |
| Myocardial infarction | No | 91.4 ± 0.4 | 85.5 ± 0.5 | 86.7 ± 0.3 |
|  | Yes | 90.7 ± 0.3 | 88.0 ± 0.2 | 86.9 ± 0.5 |
| Percutaneous coronary intervention | No | 91.1 ± 0.3 | 87.3 ± 0.2 | 87.2 ± 0.3 |
|  | Yes | 91.5 ± 0.4 | 87.6 ± 0.4 | 87.1 ± 0.7 |

**Table 3.** Discriminatory ability of PULSE–HF in ECGs with pacing and atrial fibrillation, as identified by clinician diagnoses accompanying the ECGs. These diagnoses were not available in MIMIC.

|  |  |  | AUROC |  |
| --- | --- | --- | --- | --- |
|  |  |  | Hospital 1 | Hospital 2 |
| All patients | ECGs contain pacing | No | 90.5 ± 0.4 | 87.0 ± 0.2 |
|  |  | Yes | 91.4 ± 0.8 | 87.8 ± 0.6 |
|  | ECGs contain atrial fibrillation | No | 91.4 ± 0.4 | 88.0 ± 0.2 |
|  |  | Yes | 89.1 ± 1.0 | 84.9 ± 0.6 |
| Patients with LVEF above 40% | ECGs contain pacing | No | 84.9 ± 0.7 | 81.6 ± 0.4 |
|  |  | Yes | 80.2 ± 2.1 | 80.0 ± 1.0 |
|  | ECGs contain atrial fibrillation | No | 85.5 ± 0.7 | 82.5 ± 0.4 |
|  |  | Yes | 78.8 ± 1.9 | 76.8 ± 0.9 |

**Table 4.** Cohort characteristics in patients with any LVEF > 40 after diagnosis.

|  | Hospital 1 | Hospital 2 | MIMIC |
| --- | --- | --- | --- |
| Patients, n | 39659 | 9413 | 6445 |
| Sex, % (n) | 45.4 (18,011) | 50.5 (4,751) | 52.6 (3,389) |
| Age, mean ± std | 70.5 ± 15.4 | 70.3 ± 14.6 | 73.3 ± 13.9 |
| ECGs, n | 369,985 | 78,519 | 48,354 |
| Prior 1-year LVEF support, mean | 1.09 | 1.11 | 0.99 |
| LVEF measurements, n | 159,992 | 27,280 | 17,548 |
| LVEF ≤ 40 (HFrEF), % (n) | 12.8 (20,461) | 17.2 (4,695) | 17.0 (2,987) |
| LVEF 41-49 (HFmrEF), % (n) | 10.4 (16,637) | 9.1 (2,488) | 8.8 (1,545) |
| LVEF ≥ 50 (HFpEF), % (n) | 76.8 (122,894) | 73.7 (20,097) | 74.2 (13,016) |
| Diabetes mellitus, % (n) | 42.7 (16,946) | 39.1 (3,681) | 38.9 (2,509) |
| Hypertension, % (n) | 85.2 (33,788) | 69.0 (6,494) | 62.7 (4,040) |
| Atherosclerotic heart disease, % (n) | 65.3 (25,899) | 56.3 (5,302) | 51.9 (3,344) |
| Chronic obstructive pulmonary disease, % (n) | 14.3 (5,670) | 7.2 (680) | 5.3 (342) |
| Atrial fibrillation, % (n) | 53.4 (21,176) | 48.2 (4,536) | 51.4 (3,313) |
| Coronary artery bypass graft, % (n) | 7.5 (2,982) | 3.2 (302) | 5.9 (383) |
| Percutaneous coronary intervention, % (n) | 26.2 (10,373) | 20.1 (1,889) | 10.9 (704) |
| Myocardial infarction, % (n) | 40.8 (16,169) | 53.1 (5,003) | 23.1 (1,486) |
| ACEi/ARB, % (n) | 64.6 (25,619) | 49.5 (4,655) | 54.1 (3,488) |
| ARNi, % (n) | 9.3 (3,673) | 6.1 (575) | 8.1 (519) |
| Beta blockers, % (n) | 79.2 (31,409) | 66.6 (6,267) | 83.2 (5,363) |
| MRA, % (n) | 16.3 (6,461) | 8.6 (813) | 15.8 (1,019) |
| Diuretic, % (n) | 76.3 (30,277) | 76.9 (7,240) | 85.5 (5,509) |

### Forecasting worsening LVEF in adults with baseline LVEF above 40%

To determine how well PULSE-HF can forecast worsening LVEF, the subgroup of patients where the most recent LVEF was above 40% was evaluated. PULSE-HF exhibited strong discriminative performance with AUROCs of 81.6-86.3% (Figure 2A) and AUPRCs of 56.8-67.7% (Figure 2B). When using ECGs alone, without access to LVEF history, PULSE-HF maintained AUROCs of 75.4-80.2% and AUPRCs of 42.6-59.4%. For comparison, logistic regression models achieved AUROCs of 61.3-63.7% when using ECG features only, and AUROCs 66.7-71.9% when using ECG and LVEF history features. In addition, the discriminatory ability remained consistently high across a range of subgroups with different comorbidities and regardless of medical therapy (Table 5). PULSE-HF also demonstrated consistent sensitivity–specificity tradeoffs across all three institutions, with specificity ranging from 75.3-83.9% at 70% train sensitivity, 60.3-75.2% at 80% train sensitivity, and 34.0-58.4% at 90% train sensitivity (Figure 2C). For each test dataset, sensitivities corresponding to train sensitivity thresholds are listed in Table S11.

**Figure 2.**
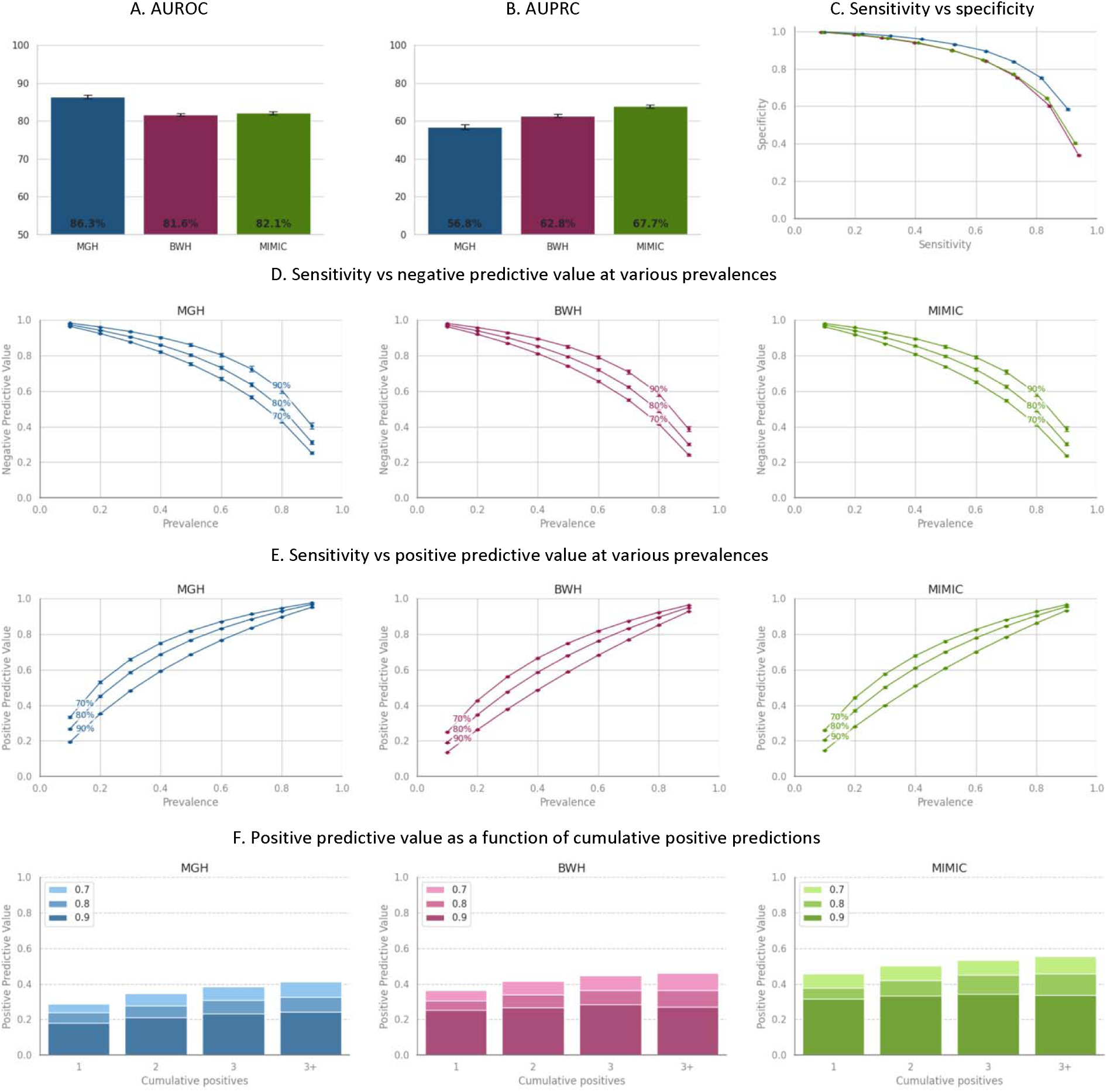
PULSE-HF performance in patients who have a baseline LVEF above 40%. Results are shown for three institutions: the internal test set at Hospital 1, and external validation at Hospital 2 and MIMIC. (A) Area under the receiver operating characteristic curve (AUROC). (B) Area under the precision-recall curve (AUPRC). (C) Sensitivity vs. specificity trade-off curves. (D) Sensitivity vs. negative predictive value (NPV) at various assumed prevalences of one-year incidence of LVEF decline below 40%. (E) Sensitivity vs. positive predictive value (PPV) at different assumed prevalences of one-year incidence of LVEF decline below 40%. (F) PPV as a function of cumulative positive predictions, demonstrating that multiple positive predictions improve PULSE–HF’s risk stratification.

**Table 5.** Discriminatory ability for forecasting worsening LVEF in patients with LVEF > 40, stratified by important clinical variables.

| Subgroup |  | AUROC |  |  |
| --- | --- | --- | --- | --- |
|  |  | Hospital 1 | Hospital 2 | MIMIC |
| All patients |  | 84.0 ± 0.5 | 81.6 ± 0.3 | 82.1 ± 0.4 |
| Average annual hospitalizations | < 1 | 84.2 ± 0.6 | 81.7 ± 0.4 | 81.0 ± 0.6 |
|  | ≥ 1 | 82.6 ± 0.9 | 80.4 ± 0.7 | 82.8 ± 0.6 |
| Diabetes mellitus | No | 83.6 ± 0.6 | 82.2 ± 0.5 | 81.9 ± 0.5 |
|  | Yes | 84.5 ± 0.7 | 80.7 ± 0.5 | 82.4 ± 0.6 |
| Hypertension | No | 82.8 ± 1.2 | 82.1 ± 0.6 | 81.8 ± 0.6 |
|  | Yes | 83.9 ± 0.6 | 80.7 ± 0.4 | 82.2 ± 0.5 |
| Atherosclerotic heart disease | No | 84.1 ± 1.0 | 81.6 ± 0.6 | 80.7 ± 0.7 |
|  | Yes | 83.9 ± 0.6 | 81.2 ± 0.4 | 82.2 ± 0.5 |
| Chronic obstructive pulmonary disease | No | 84.3 ± 0.5 | 81.4 ± 0.4 | 82.1 ± 0.4 |
|  | Yes | 81.3 ± 1.5 | 83.8 ± 1.4 | 82.6 ± 2.3 |
| Coronary artery bypass graft | No | 84.4 ± 0.5 | 81.5 ± 0.4 | 82.2 ± 0.4 |
|  | Yes | 79.8 ± 1.5 | 84.0 ± 1.8 | 80.5 ± 1.6 |
| Atrial fibrillation | No | 86.7 ± 0.6 | 83.7 ± 0.5 | 82.6 ± 0.6 |
|  | Yes | 81.6 ± 0.7 | 79.3 ± 0.5 | 81.7 ± 0.6 |
| ACEi/ARB | No | 85.6 ± 0.7 | 81.3 ± 1.1 | 81.3 ± 0.7 |
|  | Yes | 83.3 ± 0.6 | 81.5 ± 0.4 | 82.0 ± 0.5 |
| ARNi | No | 83.7 ± 0.5 | 82.5 ± 0.5 | 81.9 ± 0.4 |
|  | Yes | 87.2 ± 1.3 | 80.7 ± 0.5 | 84.3 ± 1.2 |
| MRA | No | 84.0 ± 0.5 | 81.5 ± 0.4 | 80.4 ± 0.5 |
|  | Yes | 84.3 ± 1.2 | 83.3 ± 1.5 | 87.0 ± 0.7 |
| Beta blockers | No | 85.4 ± 1.0 | 81.4 ± 0.4 | 83.5 ± 1.2 |
|  | Yes | 83.7 ± 0.5 | 83.1 ± 1.1 | 81.7 ± 0.4 |
| Diuretics | No | 82.2 ± 1.0 | 83.1 ± 0.6 | 78.2 ± 1.1 |
|  | Yes | 84.4 ± 0.5 | 80.5 ± 0.4 | 82.8 ± 0.4 |
| Myocardial infarction | No | 85.1 ± 0.7 | 81.4 ± 0.6 | 81.8 ± 0.5 |
|  | Yes | 82.2 ± 0.6 | 81.5 ± 0.4 | 80.7 ± 0.7 |
| Percutaneous coronary intervention | No | 84.4 ± 0.6 | 81.2 ± 0.4 | 81.7 ± 0.4 |
|  | Yes | 83.1 ± 0.8 | 81.9 ± 0.7 | 82.3 ± 0.9 |

Assuming a 10% incidence of worsening LVEF, NPVs and PPVs for PULSE-HF were computed as a function of model sensitivity. Calculated NPVs were 96.2-96.5% at 70% train sensitivity, 97.2-97.4% at 80% train sensitivity, and 98.1-98.2% at 90% train sensitivity (Figure 2D). Corresponding PPVs range between 24.9-33.3% at 70% train sensitivity, 19.1-26.8% at 80% train sensitivity, and 13.7-19.5% at 90% train sensitivity (Figure 2E).

Across various sensitivity thresholds, PPV increased consistently as positive predictions accumulated (Figure 2F). Among instances with at least three predictions within the two years following an ECG measurement, instances with only one positive prediction resulted in PPVs of 28.9%, 36.4%, and 45.6% at 70% train sensitivity in Hospital 1, Hospital 2, and MIMIC. With three positive predictions in that time, PPVs improved to 38.4%, 44.8%, and 53.3%, respectively.

### Forecasting worsening LVEF in adults with baseline LVEF above 40% using only lead I

Across all twelve single-lead models, discriminatory ability in forecasting LVEF below 40% remained performant (Figure S4). To further evaluate whether a single lead model could forecast worsening LVEF, the version of PULSE-HF trained using lead I alone, PULSE-HFL1, was evaluated. Lead I was chosen since most wearable and pocket devices equipped with ECG sensors record this lead (20,21). For the task of forecasting LVEF deterioration among patients with a baseline LVEF above 40%, the lead I model maintained its discriminative ability relative to the full 12-lead model, achieving AUROCs of 79.4-84.5% (Figure 3A) and AUPRCs of 51.0-63.3% (Figure 3B). When using ECGs alone, without access to LVEF history features, PULSE-HFL1 achieved AUROCs of 72.0-75.2% and AUPRCs of 33-51.9%. Sensitivity-specificity tradeoffs remained stable, with specificity ranging from 68.0-81.0% at 70% train sensitivity, 54.2-72.4% at 80% train sensitivity, and 26.5-50.7% at 90% train sensitivity (Figure 3C). At a 10% prevalence of worsening per year, NPVs ranged from 95.8-96.5% at 70% train sensitivity, 96.6-97.2% at 80% train sensitivity, and 97.9-98.3% at 90% train sensitivity (Figure 3D). PPVs were 21.3-29.8% at 70% train sensitivity, 17.3-24.6% at 80% train sensitivity, and 12.6-17.2% at 90% train sensitivity (Figure 3E). For each test dataset, sensitivities corresponding to train sensitivity thresholds are listed in Table S11.

**Figure 3.**
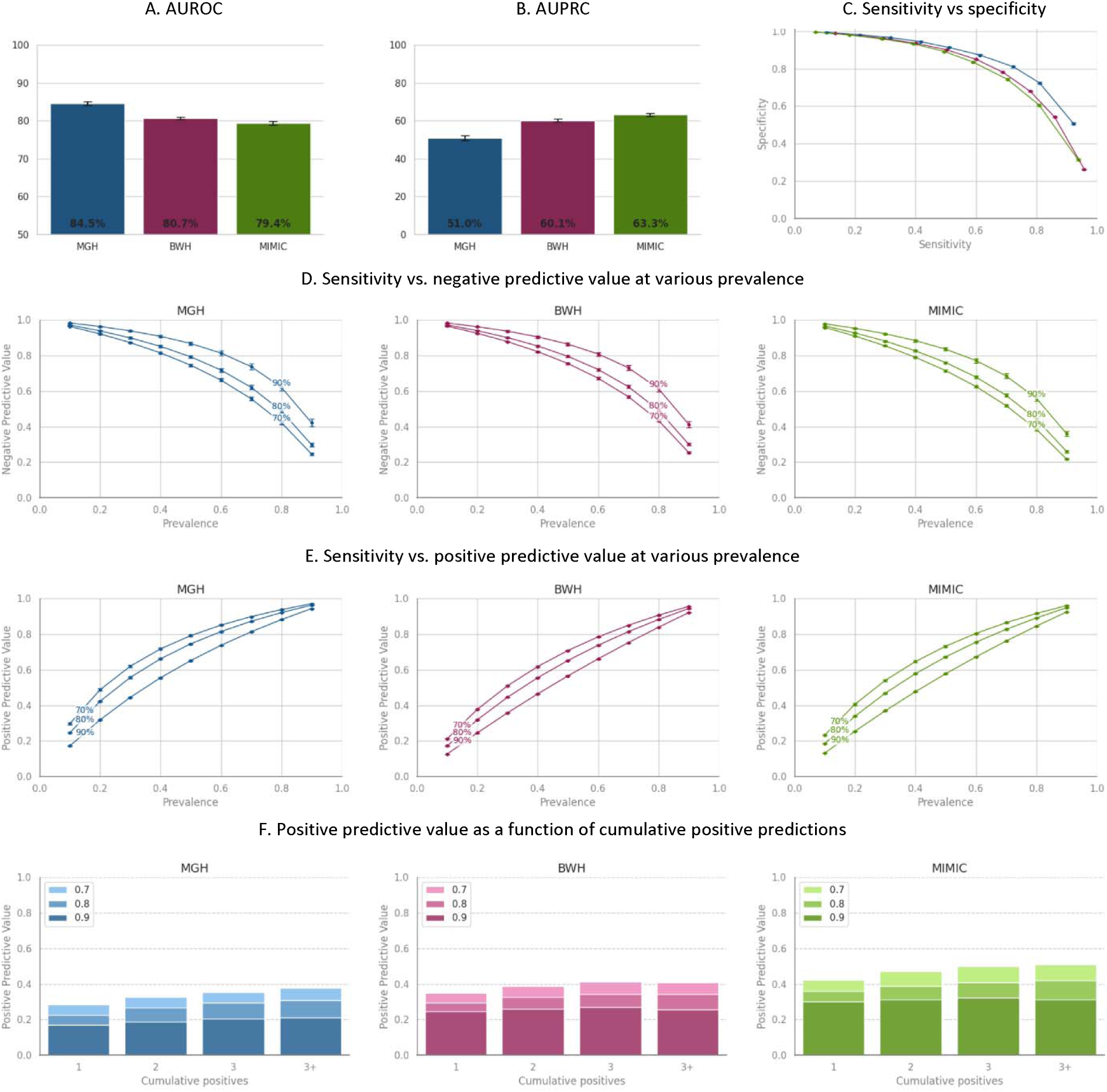
PULSE-HF Lead 1 model performance in patients who have a baseline LVEF above 40%. Results are shown for three hospitals. (A) Area under the receiver operating characteristic curve (AUROC). (B) Area under the precision-recall curve (AUPRC). (C) Sensitivity vs. specificity trade-off curves. (D) Sensitivity vs. negative predictive value (NPV) at various assumed prevalences of one-year incidence of LVEF decline below 40%. (E) Sensitivity vs. positive predictive value (PPV) at different assumed prevalences of one-year incidence of LVEF decline below 40%. (F) PPV as a function of cumulative positive predictions, demonstrating that multiple positive predictions improve PULSE–HF’s risk stratification.

PULSE-HFL1 achieved PPVs of 28.5%, 35.1%, and 42.4% at 70% train sensitivity in Hospital 1, Hospital 2, and MIMIC, with only one positive prediction over the previous two years, among instances with at least three predictions. With three positive predictions, PPVs improved to 35.3%, 41.4%, and 49.8%, respectively.

### Determinants of PULSE-HF performance

To investigate the relative importance of ECG and LVEF features for PULSE-HF1 performance, saliency analysis was performed, which identified the portions of the input that most influenced the model prediction. This saliency analysis revealed that PULSE-HFL1 predictions were primarily impacted by ECG features, particularly features occurring during diastole, as saliency values for ECG-based features were one order of magnitude larger than those for LVEF history (Figures 4A & B). Moreover, the highest saliency values were associated with the portions of the ECG signal originating from the 200ms preceding the subsequent R-wave – corresponding to late ventricular diastole (Figure 4A). Example ECG signals with superimposed saliency maps are shown in Figure 5. In each case, the highest saliency values occur between the T-wave and the onset of the QRS complex.

**Figure 4.**
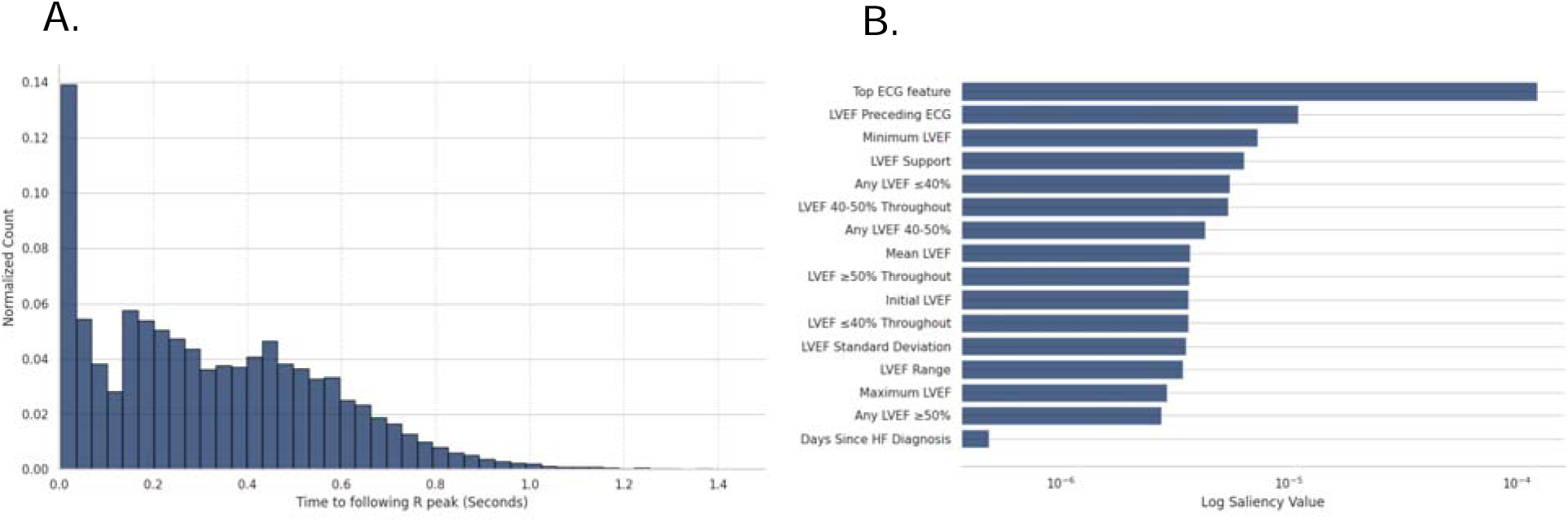
Determinants of PULSE–HFL1 performance (A) Distribution of the time difference between the most salient ECG feature used by PULSE–HF and the R-peak that immediately follows this feature. (B) Mean saliency values of LVEF history features and the ECG feature with the highest saliency.

**Figure 5.**
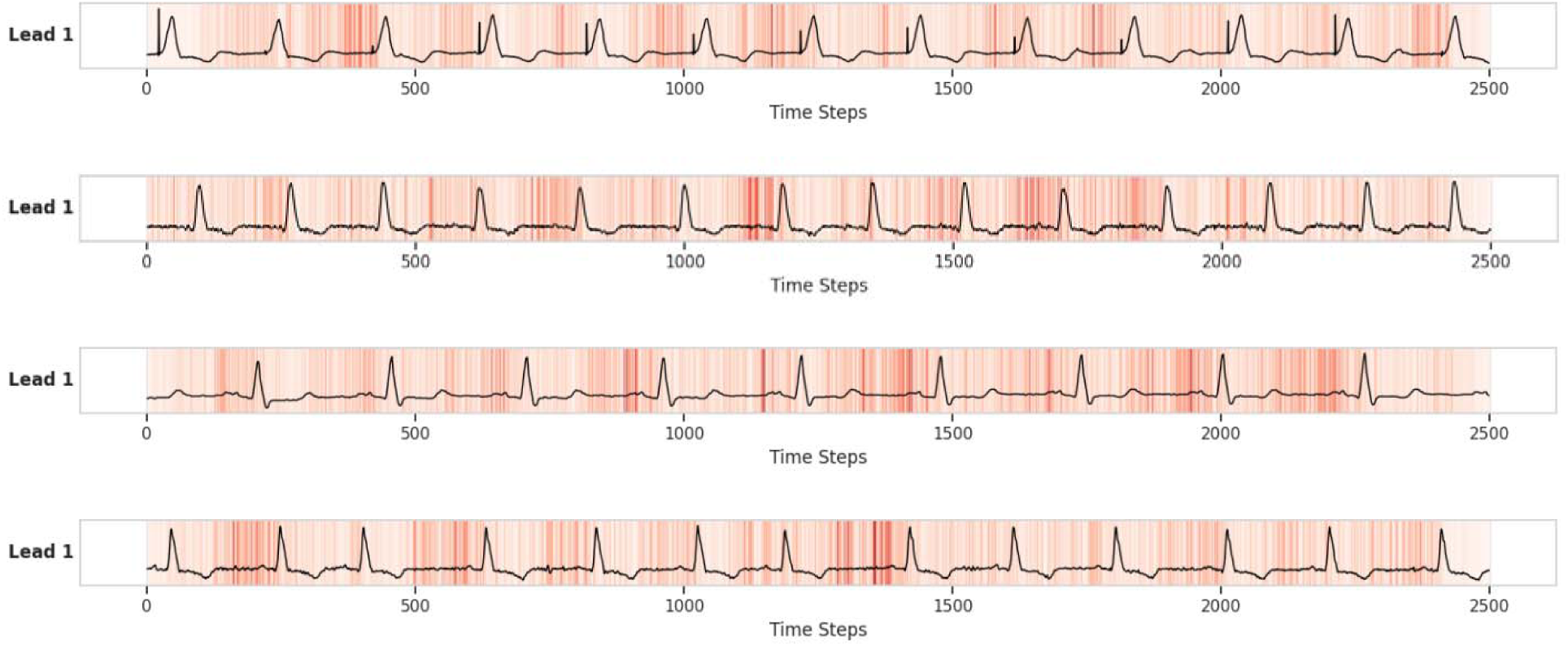
Saliency maps of PULSE-HFL1. The darker colors represent larger gradients.

## Discussion

PULSE–HF leverages 12-lead ECGs and LVEF history to predict whether a patient’s LVEF will be below 40% within one year after the index ECG. The model is designed specifically for prognostication in patients who have a prior diagnosis of HF and was validated across three retrospective datasets of patients who have a prior diagnosis of HF. Among these retrospective cohorts, the discriminatory ability was high (AUROCs 87.5-91.4%) and remained high across subgroups with different comorbidities and medical therapies. In the baseline logistic regression models, the AUROCs are lower than those of PULSE-HF, achieving merely ∼69-70% using ECGs alone and ∼74-78% when also using LVEF history. The performance gap highlights the benefits of leveraging deep learning for this forecasting task.

In patients who have an LVEF below 40%, PULSE-HF identifies individuals who will have persistence or worsening of their LV systolic function. Such patients may benefit from closer monitoring that could enable earlier evaluation for advanced therapies (3). Arguably, however, a more important application for PULSE–HF is in identifying patients who have an initial LVEF over 40% and will have a decline below 40% in their LVEF in the near future. In these patients, PULSE-HF can discriminate between patients who will have a subsequent decline in their LVEF versus those who will not (AUROCs 81.6-86.3%). The high discriminatory ability is again preserved over all three clinical cohorts and across subgroups with different comorbidities. Prior attempts to forecast worsening LVEF using structured EHR data have shown limited success, with AUROCs ranging from 54% to 68% (12) and 79% for a shorter future time window of six months (15), underscoring the importance of ECGs in capturing subclinical markers of systolic dysfunction. Moreover, a logistic regression model trained using ECG features and information about the LVEF history achieves AUROCs of 66.7-71.9%. Assuming an underlying incidence for LVEF worsening of 10% per year, and a 70% train sensitivity, PULSE–HF demonstrated modest PPVs (24.9-33.3%) and high NPVs (96.2-96.5%) across all three institutions. Patients who have multiple positive predictions have higher PPVs (38.4-53.3%). These data suggest that PULSE-HF provides a robust approach for identifying patients at low risk of future LVEF decline.

The ability to predict LVEF decline using single lead ECGs with a strong performance comparable to the 12-lead model would facilitate the identification of high-risk patients using wearable and/or pocket ECG monitors, which most often record ECG signals using a single lead (20,21). This highlights the potential for integrating PULSE–HF into remote monitoring devices, which would allow for continuous risk assessment outside of traditional healthcare environments.

Current AHA guidelines recommend reassessment only when it would meaningfully alter clinical management (22). Patients who are deemed to be at low risk of adverse outcomes may therefore receive fewer follow-up echocardiograms relative to those who are deemed to be at high risk. A clinician’s ability to detect LVEF decline promptly is limited by their ability to risk stratify based on clinical signs, symptoms, and the EHR. EHR data can be challenging to process due to high missingness and noise; however, obtaining 12-lead ECGs has become a routine part of annual visits to a cardiovascular clinic. By leveraging ECGs, PULSE-HF provides a low-cost annual screening method to longitudinally identify patients with HF at high risk of LVEF decline. Patients identified as high risk by PULSE–HF could be prioritized for follow-up echocardiography, thereby enabling the timely identification of HFrEF. By detecting left ventricular dysfunction sooner, clinicians could initiate GDMT earlier, which has been shown to improve survival, reduce HF hospitalizations, slow disease progression, and enhance quality of life for patients with HFrEF (23).

While deep learning models provide a platform for building performant models using ECG data, it is challenging to understand precisely what these models have learned and how they arrive at decisions. Insights into what aspects of the input signal the model tends to focus on when arriving at a decision can sometimes be garnered from a saliency analysis, which strives to understand what input features influence model predictions the most (17,18,24). In PULSE-HFL1, the model tends to focus on regions of the ECG signal within 200ms before the subsequent R-wave – the time corresponding to end diastole. This suggests a potential link between impairments in left ventricular filling and future declines in systolic function. Although PULSE-HFL1 integrates both ECG and LVEF history, decisions were largely driven by ECG-derived features. Saliency values for ECG-based features are one order of magnitude larger than those for LVEF history, indicating that subtle temporal variations in the ECG signal provide meaningful prognostic information beyond what LVEF alone can capture. While LVEF features are significantly less influential than ECG features, the LVEF measurement taken immediately before the ECG is the most relevant.

Our study has limitations. Our method is retrospectively evaluated on three clinical cohorts, corresponding to observational datasets. As such, the timing in which echocardiograms were obtained was determined by subjective clinical assessment and clinical practice, which can vary at different institutions, and by patient adherence in completing testing when ordered. Differences in care practices and resource availability at other sites could introduce biases that warrant further investigation. Furthermore, our cohort for training included patients who had at least one LVEF within a year following an ECG, so a label could be defined. Hence, patients who have rapid clinical deterioration, resulting in death within a year after an index ECG but prior to any subsequent LVEF measurement, were excluded from our analysis. Prospective evaluation is necessary to assess the utility of PULSE–HF in guiding HF management decisions, optimizing prediction windows, and integrating the model into clinical practice. While the single-lead models developed from 12-lead data are highly effective, it remains to be seen whether the same performance can be achieved with real-world, potentially noisier wearable ECG data.

PULSE–HF is a performant AI-based approach for forecasting worsening systolic function in patients with HF and leverages a widely available test – the ECG – for risk stratification. PULSE–HF has the potential to improve HF outcomes by enabling timely interventions before irreversible cardiac deterioration occurs.

## Data Availability

All data produced in the present study are available upon reasonable request to the authors and approval by the IRB

## Data sharing statement

Raw ECG and EHR data can be shared pending IRB approval at the relevant hospital from which the data was collected. Please reach out to the corresponding authors for more information. PULSE–HF model weights and development code are freely available at https://github.com/mit-ccrg/PULSE-HF.

## Acknowledgements

We would like to thank Leo Anthony Celi, Takeshi Tohyama, and Brian Gow for their help with accessing data from the MIMIC database.

