## Supplemental Information for "Forecasting left ventricular systolic dysfunction in heart failure with artificial intelligence"

**Supplementary Materials**

**Figure S1A.**

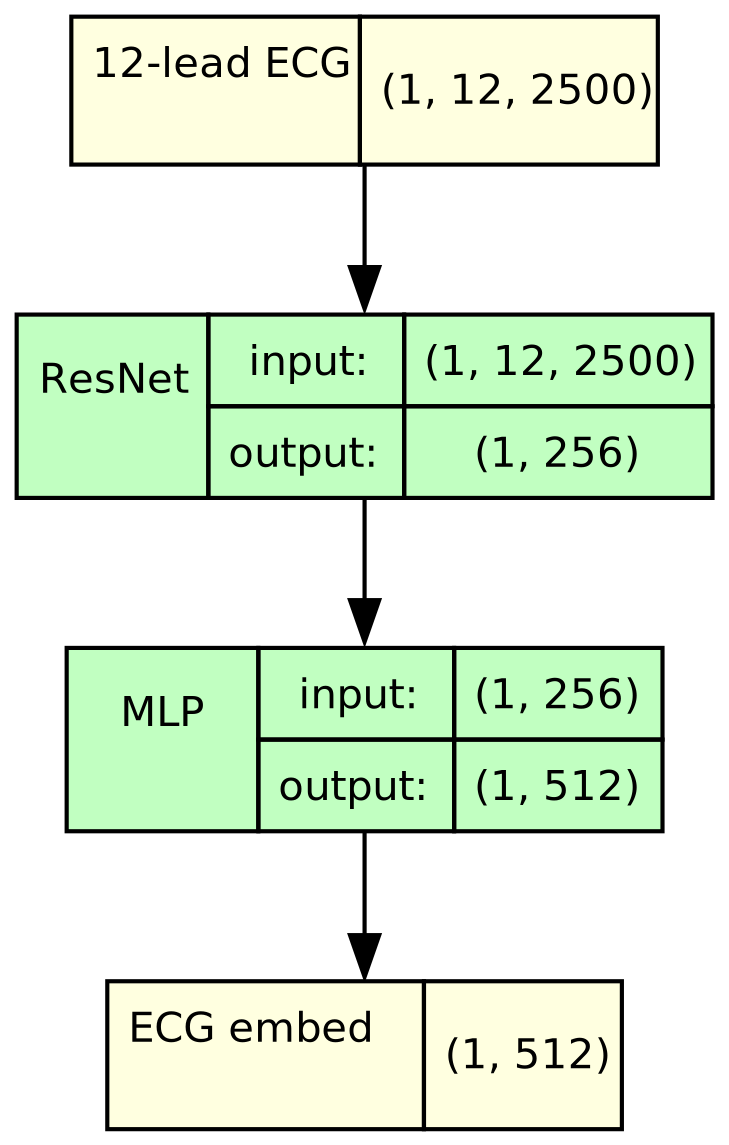

PULSE–HF ECG encoder

**Figure S1B.** PULSE-HF ECG encoder design

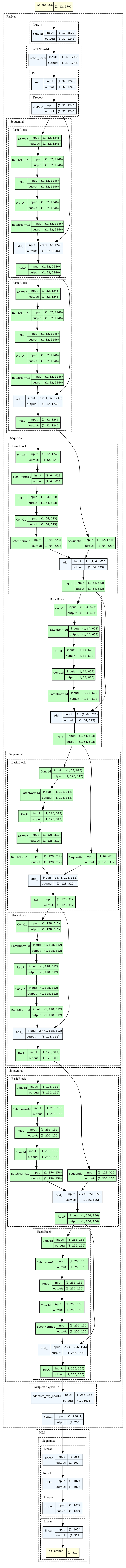

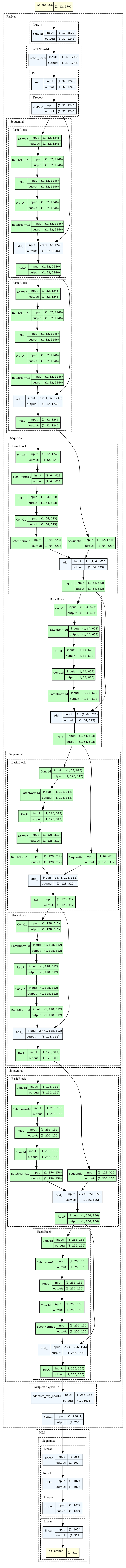

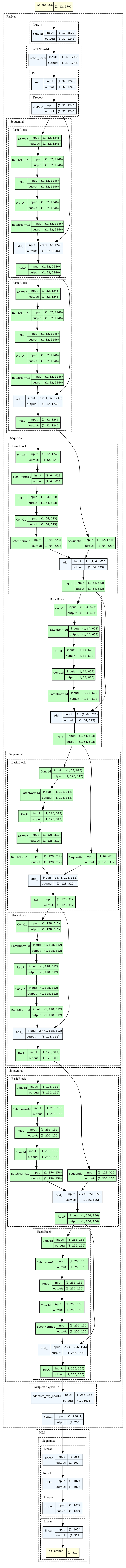

**Figure S2.**

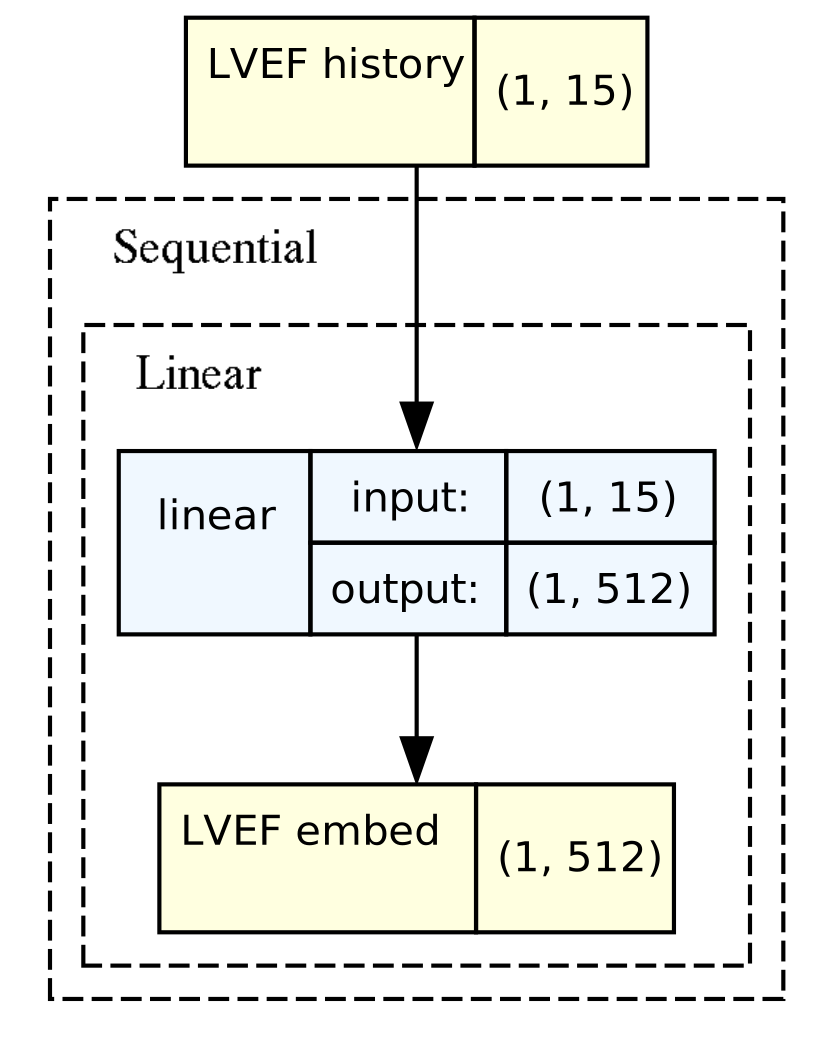

PULSE–HF LVEF history encoder

**Figure S3**. PULSE–HF prediction module.

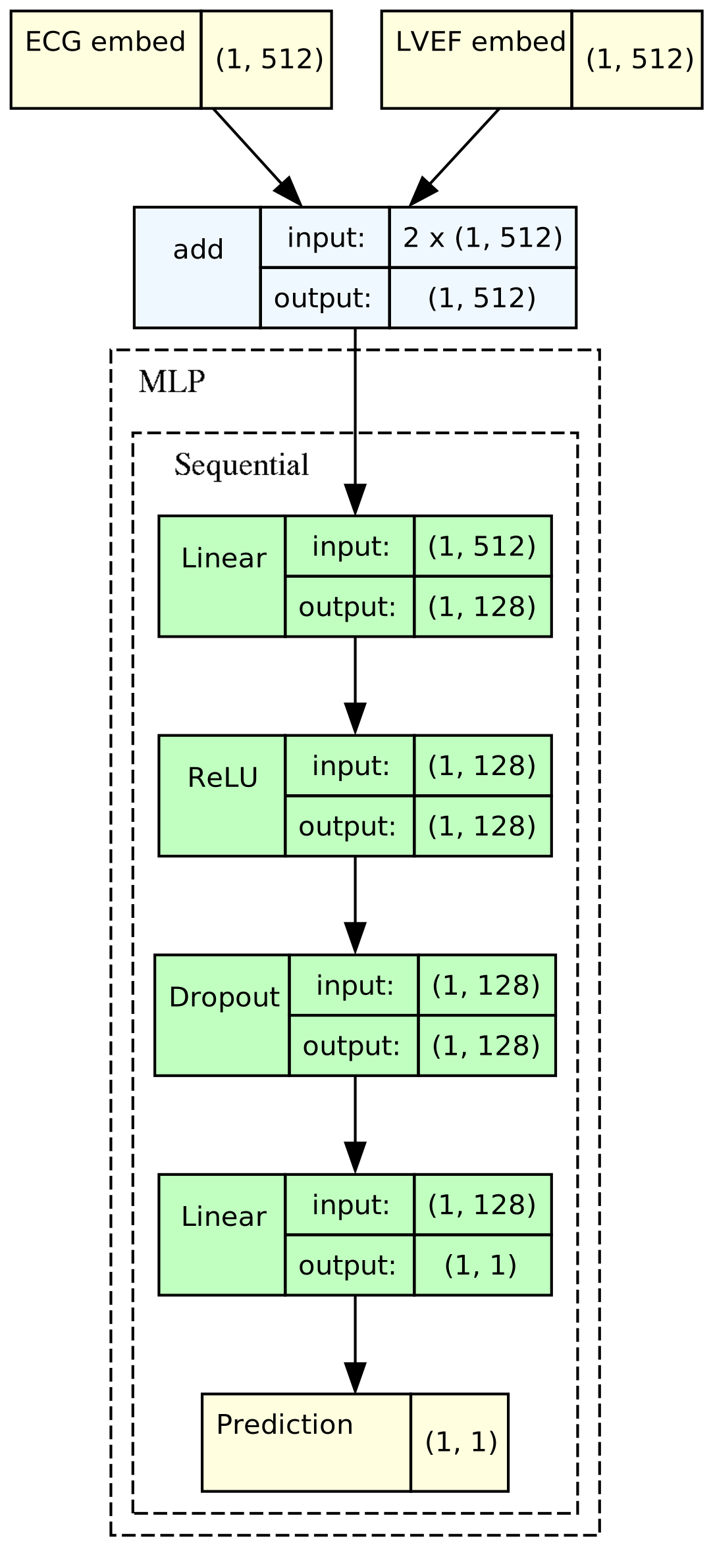

**Figure S4**. AUROC of single lead models in forecasting LVEF ≤ 40% among all patients. Horizontal baseline is AUROC for 12-lead PULSE–HF.

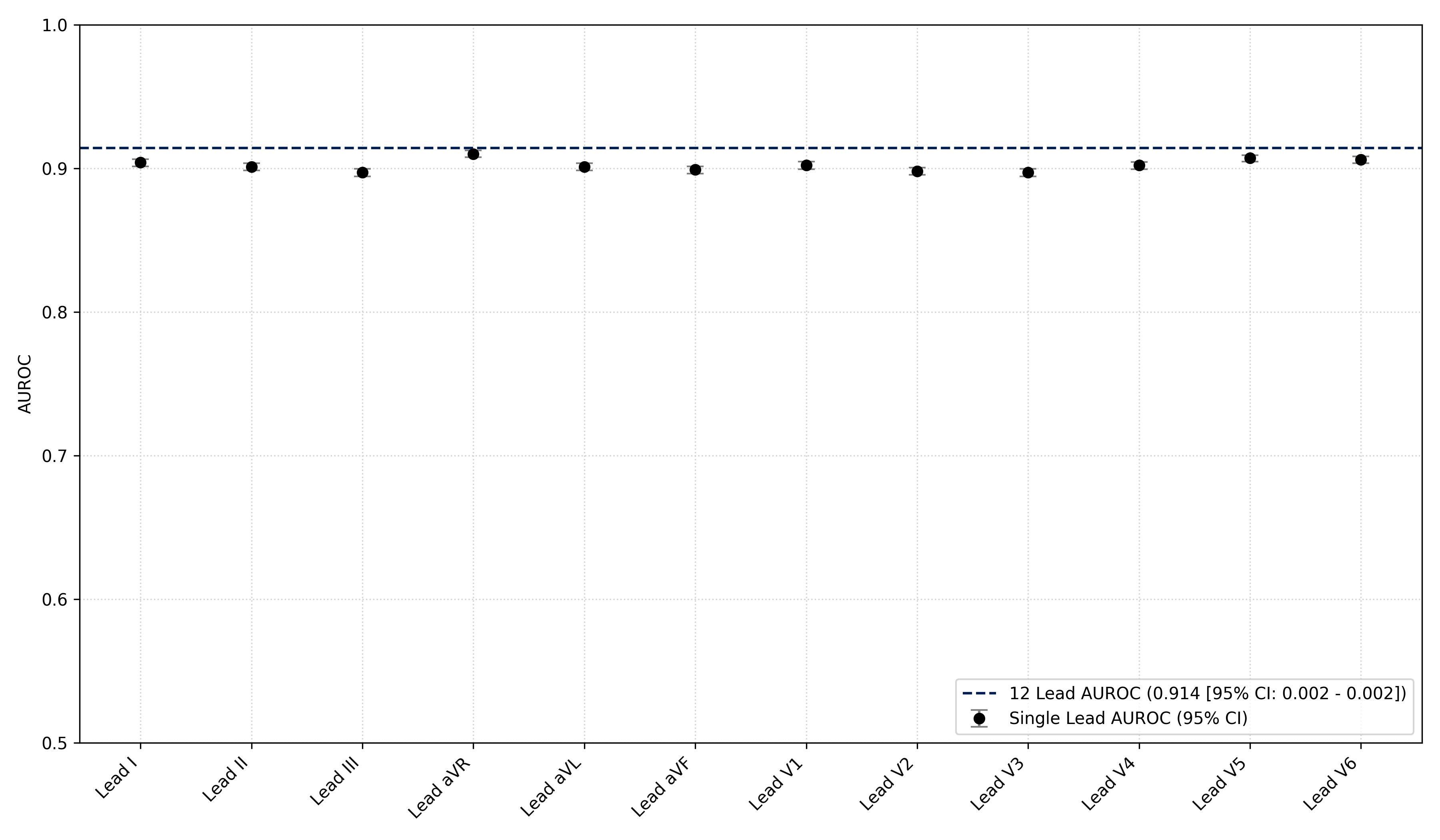

**Figure S5**. PULSE-HF performance in patients who have a baseline LVEF in HFrEF (≤ 40%). Results are shown for three hospitals. (A) Area under the receiver operating characteristic curve (AUROC). (B) Area under the precision-recall curve (AUPRC). (C) Sensitivity vs. specificity trade-off curves. (D) Sensitivity vs. negative predictive value (NPV) at various assumed prevalences of one-year incidence of LVEF decline below 40%. (E) Sensitivity vs. positive predictive value (PPV) at different assumed prevalences of one-year incidence of LVEF decline below 40%.

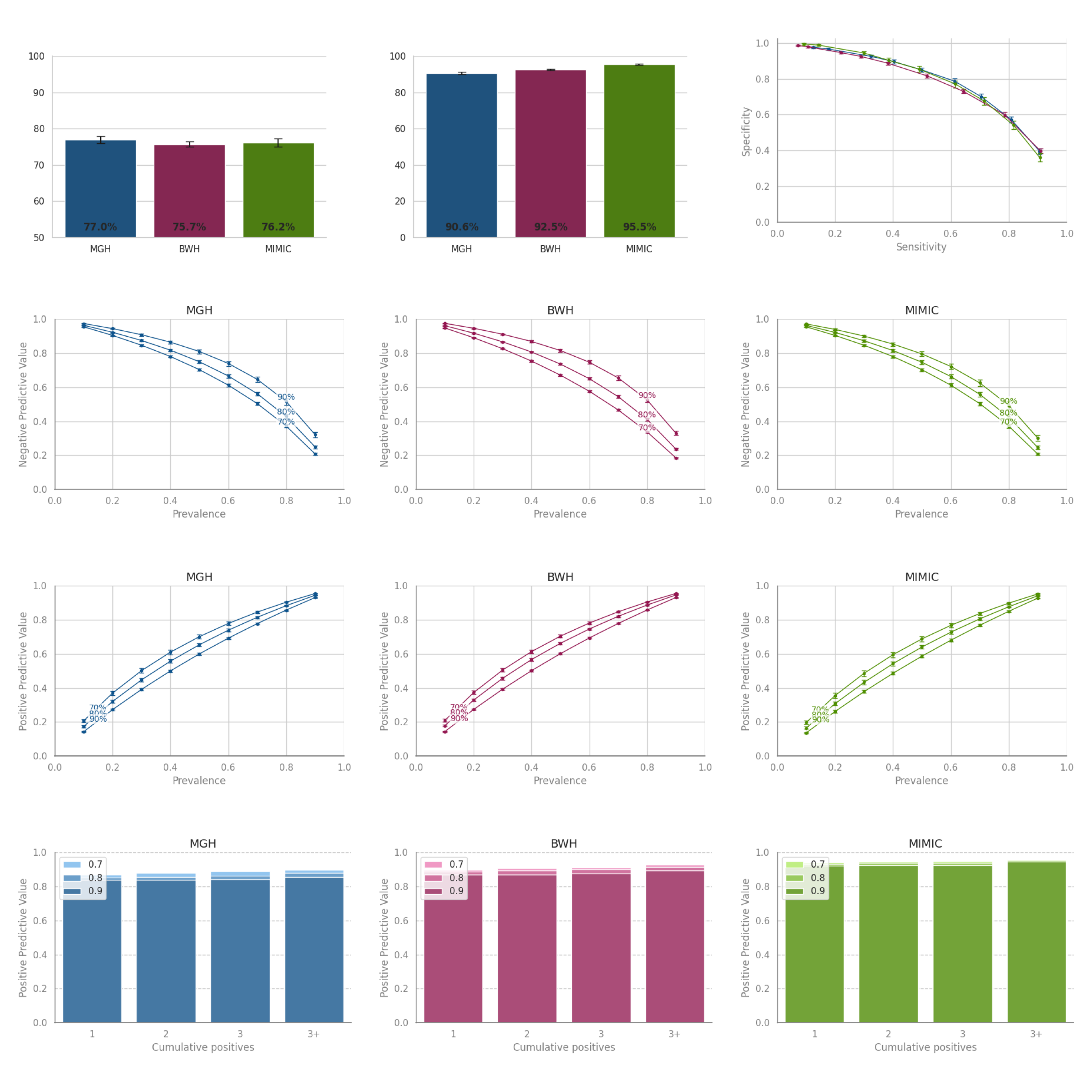

A. AUROC

B. AUPRC

C. Sensitivity vs specificity

D. Sensitivity vs negative predictive value at various prevalences

E. Sensitivity vs positive predictive value at various prevalences

F. Positive predictive value as a function of cumulative positive predictions

**Figure S6**. PULSE-HF performance in patients who have a baseline LVEF in HFrEF (≤ 40%) to predict improvement (all LVEF above 40%) in one year after ECG. Results are shown for three hospitals. (A) Area under the receiver operating characteristic curve (AUROC). (B) Area under the precision-recall curve (AUPRC). (C) Sensitivity vs. specificity trade-off curves. (D) Sensitivity vs. negative predictive value (NPV) at various assumed prevalences of one-year incidence of LVEF decline below 40%. (E) Sensitivity vs. positive predictive value (PPV) at different assumed prevalences of one-year incidence of LVEF decline below 40%.

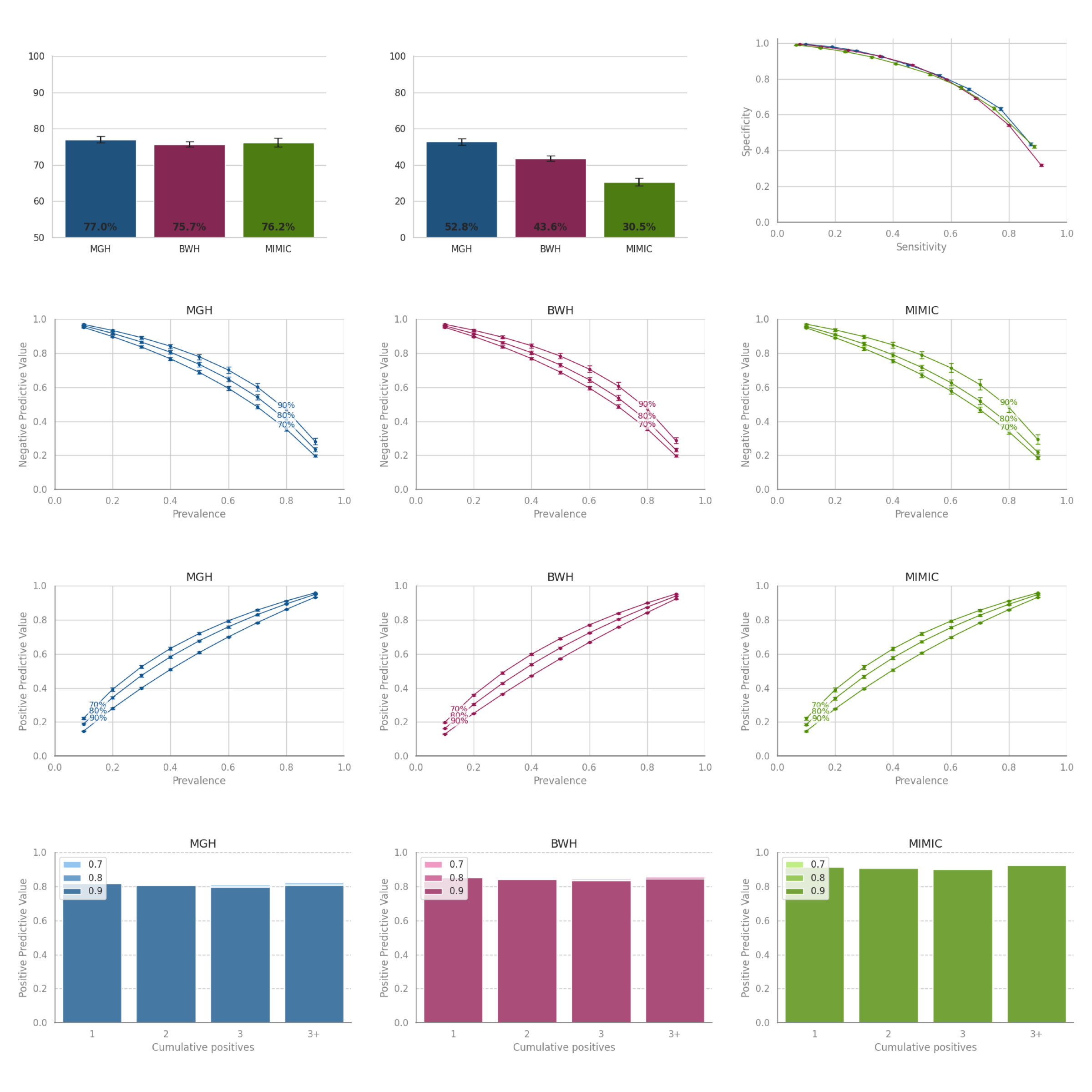

A. AUROC

B. AUPRC

C. Sensitivity vs specificity

D. Sensitivity vs negative predictive value at various prevalences

E. Sensitivity vs positive predictive value at various prevalences

F. Positive predictive value as a function of cumulative positive predictions

**Figure S7**. PULSE-HF performance in patients who have a baseline LVEF in HFpEF (≥ 50%). Results are shown for three hospitals. (A) Area under the receiver operating characteristic curve (AUROC). (B) Area under the precision-recall curve (AUPRC). (C) Sensitivity vs. specificity trade-off curves. (D) Sensitivity vs. negative predictive value (NPV) at various assumed prevalences of one-year incidence of LVEF decline below 40%. (E) Sensitivity vs. positive predictive value (PPV) at different assumed prevalences of one-year incidence of LVEF decline below 40%.

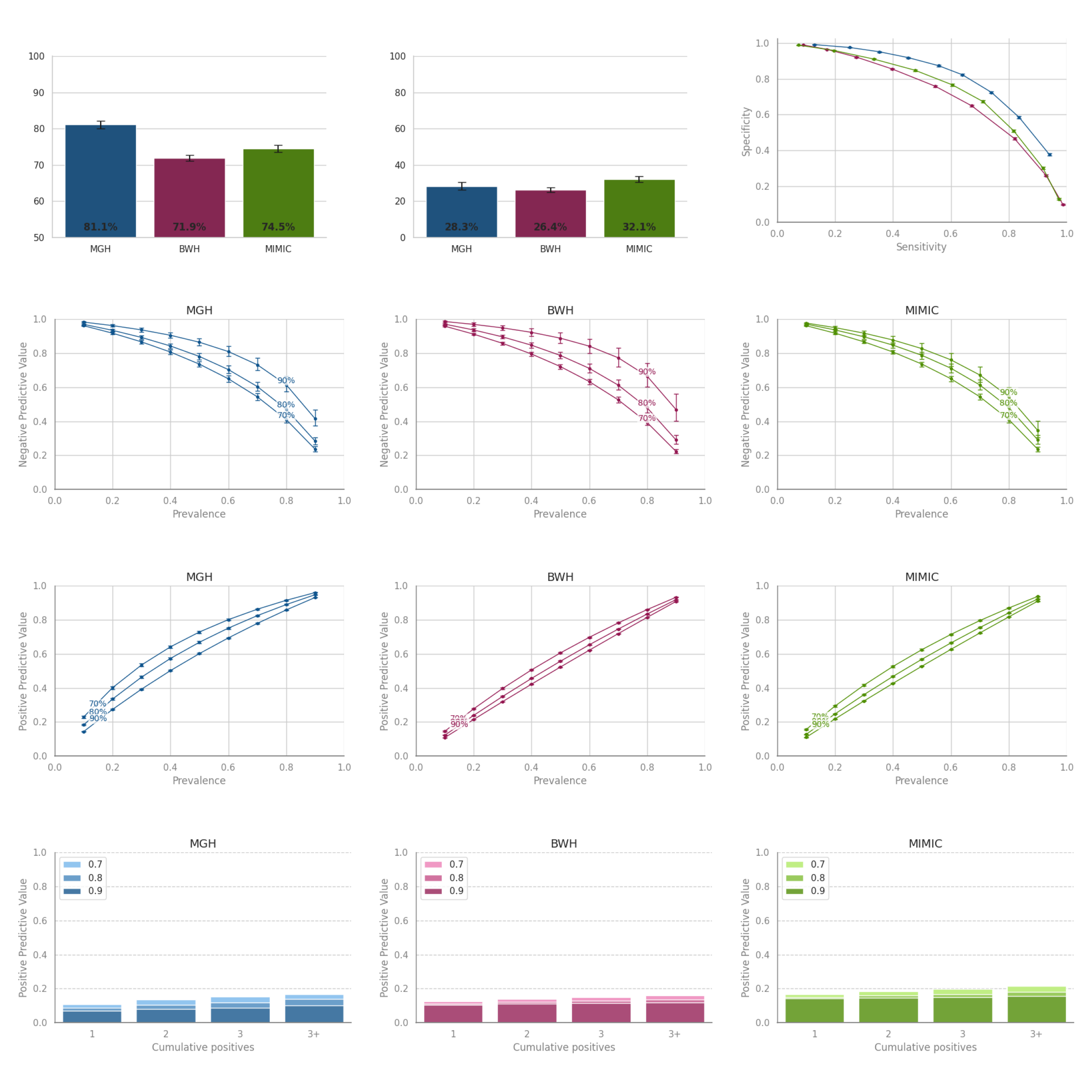

A. AUROC

B. AUPRC

C. Sensitivity vs specificity

D. Sensitivity vs negative predictive value at various prevalences

E. Sensitivity vs positive predictive value at various prevalences

F. Positive predictive value as a function of cumulative positive predictions

**Figure S8**. PULSE-HF performance in patients who have a baseline LVEF in HFmrEF (41-49%). Results are shown for three hospitals. (A) Area under the receiver operating characteristic curve (AUROC). (B) Area under the precision-recall curve (AUPRC). (C) Sensitivity vs. specificity trade-off curves. (D) Sensitivity vs. negative predictive value (NPV) at various assumed prevalences of one-year incidence of LVEF decline below 40%. (E) Sensitivity vs. positive predictive value (PPV) at different assumed prevalences of one-year incidence of LVEF decline below 40%.

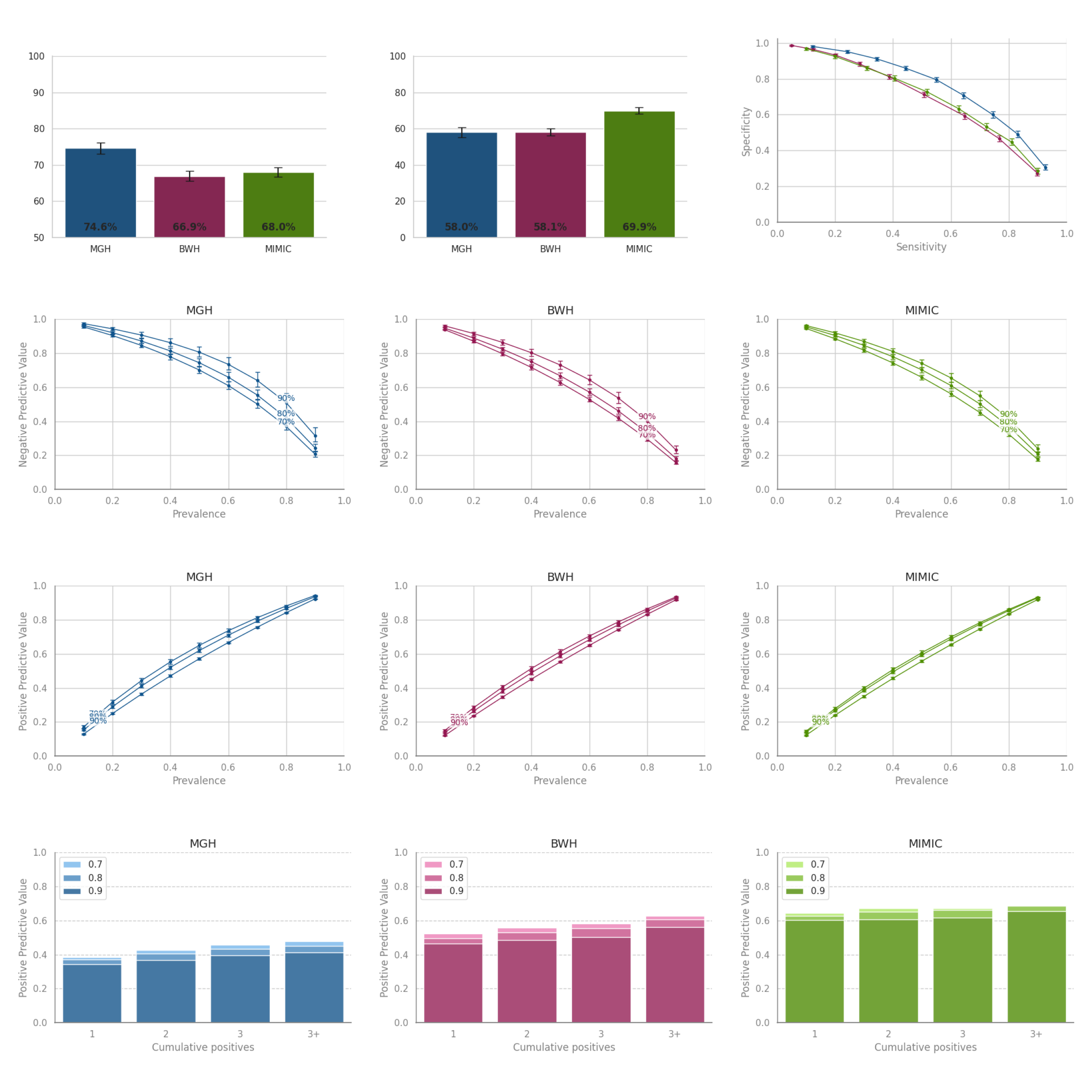

A. AUROC

B. AUPRC

C. Sensitivity vs specificity

D. Sensitivity vs negative predictive value at various prevalences

E. Sensitivity vs positive predictive value at various prevalences

F. Positive predictive value as a function of cumulative positive predictions

**Figure S9**. PULSE-HF calibration curves.

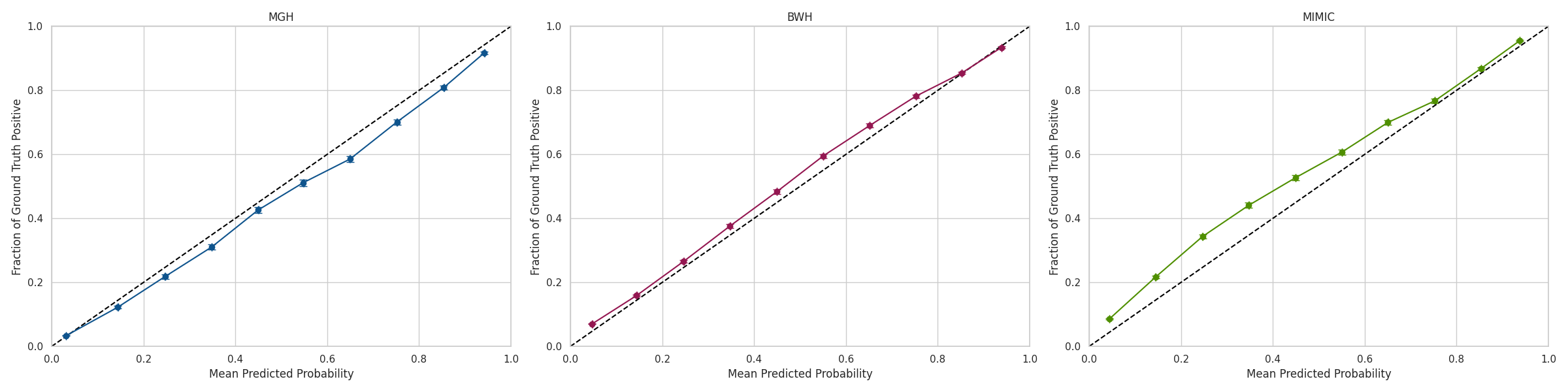

**Table S10**. ECG features used for logistic regression.

| **Feature** | **Description** |
| --- | --- |
| Atrial rate | The rate of atrial depolarization (beats per minute). |
| P-axis | The electrical axis of the P wave (in degrees), indicating the direction of atrial depolarization. |
| P-onset | The time point marking the beginning of the P wave, representing the start of atrial depolarization. |
| P-offset | The time point marking the end of the P wave, representing the completion of atrial depolarization. |
| PR-interval | The PR interval duration (milliseconds), measuring the time required for atrial depolarization. |
| Q-onset | The time point marking the beginning of the QRS complex, representing the onset of ventricular depolarization. |
| Q-offset | The time point marking the end of the QRS complex, representing the completion of ventricular depolarization. |
| QRS count | The number of detected QRS complexes (heartbeats) in the ECG segment. |
| QRS-duration | The duration (milliseconds) of the QRS complex, reflecting the time required for ventricular depolarization. |
| QT-interval | The QT interval duration (milliseconds), measuring the time from Q wave onset to T wave end (i.e., total duration of ventricular depolarization and repolarization). |
| QTc | The corrected QT interval (milliseconds), adjusted to allow comparison across heart rates. |
| T-offset | The time point marking the end of the T wave, representing the completion of ventricular repolarization. |
| Ventricular rate | The rate of ventricular depolarization (beats per minute). |

**Table S11**. PULSE-HF and PULSE-HFL1 test set sensitivity values corresponding to train sensitivities of 70%, 80%, and 90%, for patients with baseline LVEF > 40%.

| **Model** | **Site** | **Train Sensitivity=70%** | **Train Sensitivity=80%** | **Train Sensitivity=90%** |
| --- | --- | --- | --- | --- |
| PULSE-HF | MGH | 72.5 | 81.7 | 90.5 |
|  | BWH | 73.8 | 84.4 | 94.0 |
|  | MIMIC | 72.6 | 83.5 | 92.9 |
| PULSE-HFL1 | MGH | 72.4 | 81.1 | 92.3 |
|  | BWH | 77.9 | 86.1 | 95.8 |
|  | MIMIC | 70.4 | 80.9 | 93.8 |
